# Simultaneous Detection and Mutation Surveillance of SARS-CoV-2 and co-infections of multiple respiratory viruses by Rapid field-deployable sequencing

**DOI:** 10.1101/2020.06.12.20129247

**Authors:** Chongwei Bi, Gerardo Ramos-Mandujano, Sharif Hala, Jinna Xu, Sara Mfarrej, Yeteng Tian, Concepcion Rodriguez Esteban, Estrella Nuñez Delicado, Fadwa S. Alofi, Asim Khogeer, Anwar M. Hashem, Naif A.M. Almontashiri, Arnab Pain, Juan Carlos Izpisua Belmonte, Mo Li

## Abstract

Strategies for monitoring the COVID-19 infection are crucial for combating the pandemic. Here we describe a method for multiplex isothermal amplification-based sequencing and real-time analysis of multiple viral genomes. It can simultaneously detect SARS-CoV-2 and co-infecting respiratory viruses, and monitor mutations for up to 96 samples in real time. The method, termed NIRVANA for <u>N</u>anopore sequencing of <u>I</u>sothermal <u>R</u>apid <u>V</u>iral <u>A</u>mplification for <u>N</u>ear real-time <u>A</u>nalysis, showed high sensitivity and specificity for SARS-CoV-2 in 70 clinical samples. It also simultaneously detected other viral pathogens (e.g. influenza A) in clinical and municipal wastewater samples. It provides a rapid field-deployable solution of COVID-19 and cos-infection detection and surveillance of the evolution of pandemic strains.

## Introduction

The novel coronavirus disease (COVID-19) pandemic is one of the most serious challenges to public health and global economy in modern history. SARS-CoV-2 is a positive-sense RNA betacoronavirus that causes COVID-19^1^. It was identified as the pathogenic cause of an outbreak of viral pneumonia of unknown etiology in Wuhan, China, by the Chinese Center for Disease Control and Prevention (CCDC) on Jan 7, 2020^2^. Three days later, the first SARS-CoV-2 genome was released online through a collaborative effort by scientists in universities in China and Australia and Chinese public health agencies (GenBank: MN908947.3)^3^. One week after the publication of the genome the first diagnostic detection of SARS-CoV-2 using real-time reverse transcription polymerase chain reaction (rRT-PCR) was released by a group in Germany^4^.

To date, rRT-PCR assays of various designs, including one approved by the US Centers for Disease Control and Prevention (US CDC) under emergency use authorization (EUA)^5^, have remained the predominant diagnostic method for SARS-CoV-2. Although proven sensitive and specific for providing a positive or negative answer, rRT-PCR provides little information on the genomic sequence of the virus, knowledge of which is crucial for monitoring how SARS-CoV-2 is evolving and spreading and ensuring successful development of new diagnostic tests and vaccines. To this end, samples need to go through a separate workflow–typically Illumina shotgun metagenomics or targeted next-generation sequencing (NGS)^6^. Because NGS requires complicated molecular biology procedures and high-value instruments in centralized laboratories, it is performed in < 1% as many cases as rRT-PCR, evidenced by the number of genomes in the GISAID database (39,954) and confirmed global cases tallied by the WHO (6,535,354) as of June 5, 2020.

SARS-CoV-2 infections often cause symptoms similar to other respiratory viruses, thus making it challenging to distinguish co-infection especially in the flu season. Several studies have reported co-infection of SARS-CoV-2 and other respiratory viruses–respiratory syncytial virus (RSV) and influenza being the most common viral pathogens identified^7,8^, and influenza was particularly high in dead patients^9^. The detection of co-infection is potentially useful for monitoring the SARS-CoV-2 pandemic and benefiting the treatment of patients. To date, no method can rapidly diagnose multiple viral infections in a high-throughput manner. Ideally, such methods should be field-deployable to allow timely assessment of outbreaks anywhere anytime.

Both rRT-PCR and NGS are sophisticated techniques whose implementation is contingent on the availability of highly-specialized facilities, personnel and reagents. These limitations could translate into long turn-over time and inadequate access to tests even in developed countries. To address these issues, several PCR-free nucleic acid detection assays have been proposed as point-of-care replacements of rRT-PCR. Chief among them is reverse transcription coupled loop-mediated isothermal amplification (RT-LAMP^10^), which has been used for rapid detection of SARS-CoV-2 RNA^11-14^. On the sequencing front, the pocket-sized Oxford Nanopore MinION sequencer has been used for rapid pathogen identification in the field^15,16^. Because MinION offers base calling on the fly, it is an attractive platform for consolidating viral nucleic acid detection by PCR-free rapid isothermal amplification and viral mutation monitoring by sequencing. However, there are several challenges for an integrated point-of-care solution based on RT-LAMP and Nanopore sequencing. RT-LAMP requires a complex mixture of primers that increases the chance of non-specific amplification and makes it difficult to multiplex. Additionally, LAMP amplicons used for SARS-CoV-2 detection are short^12,13^. Sequencing singleplex short amplicons not only fails to take advantage of the long-read and sequencing throughput (∼10 Gb) of the MinION flow cell, it is also prone to false negative reporting due to amplification failure. To the best of our knowledge, no multiplexed isothermal amplification of SARS-CoV-2 has been reported. Nanopore sequencing has its own caveats too. Due to its relatively high basecalling error, new bioinformatics tools based on dedicated algorithms^17,18^ are also needed to accurate call the presence of viral sequences (substituting for rRT-PCR) and analyze virus mutations (substituting for NGS).

Here we developed isothermal recombinase polymerase amplification (RPA) assays to simultaneously amplify multiple regions (up to 2184 bp) of the SARS-CoV-2 genome and three respiratory viruses. This forms the basis for an integrated workflow to detect the presence of viral sequences and monitor mutations in multiple regions of the SARS-CoV-2 genome in up to 96 patients at a time (Fig. 1a). We developed a bioinformatics pipeline for on-the-fly analysis to reduce the time to answer and sequencing cost by stopping the sequencing run when data are sufficient to provide confident answers.

**Figure 1.**
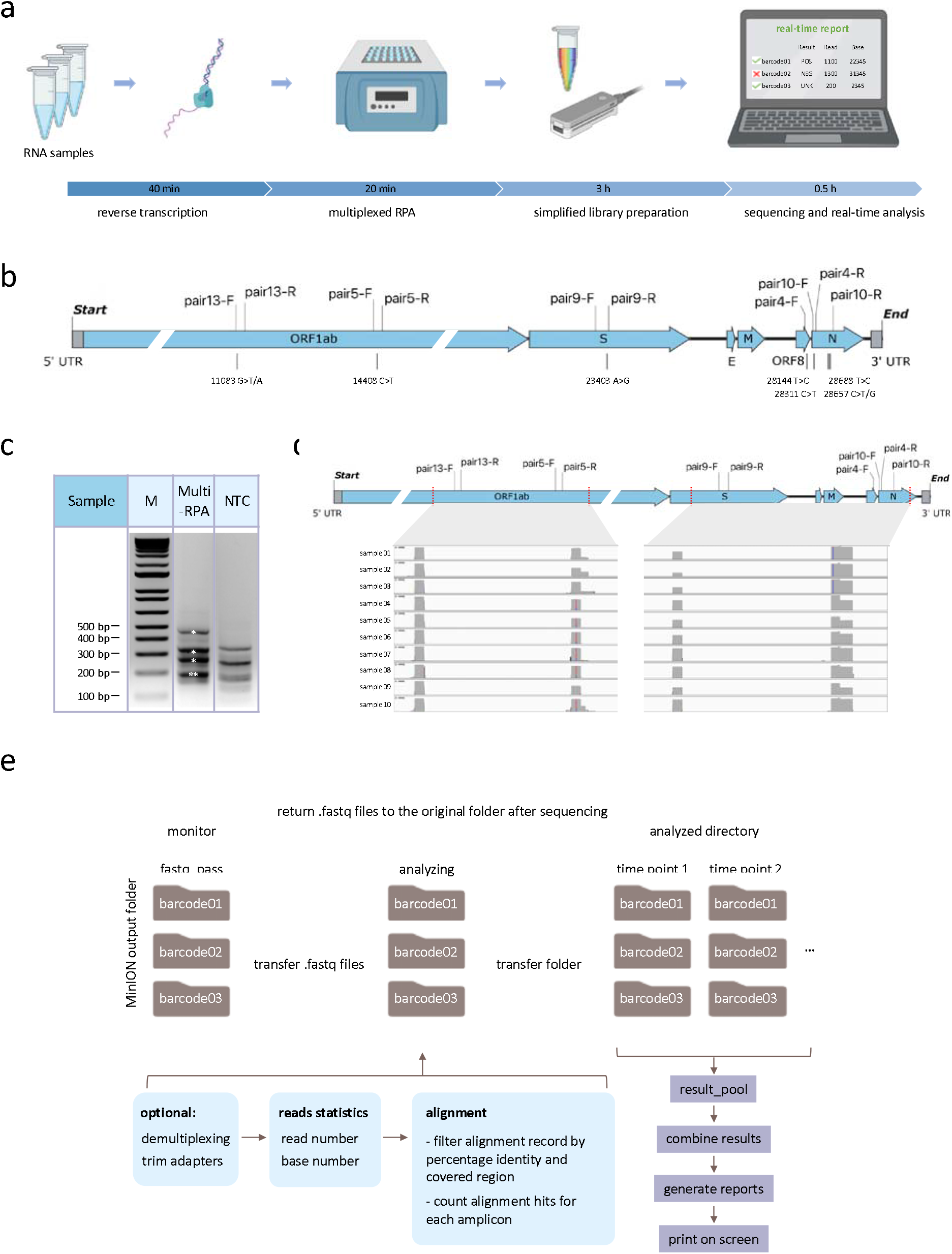
Multiplex RPA workflow for SARS-CoV-2 detection and Nanopore sequencing. **a**, Schematic representation of NIRVANA. RNA samples were subjected to reverse transcription, followed by multiplex RPA to amplify multiple regions of the SARS-CoV-2 genome. The amplicons were, purified and prepared to the Nanopore library using an optimized barcoding library preparation protocol. In the end, the sequencing was performed in the pocket-sized Nanopore MinION sequencer and sequencing results were analyzed by our algorithm termed RTNano on the fly. **b**, The RPA primers used in this study were plotted in the SARS-CoV-2 genome. The RPA amplicons are highlighted in red. The corresponding prevalent variants were labeled under the genome. **c**, Agarose gel electrophoresis results of multiplex RPA. All of the five amplicons were shown in the gel with correct size (asterisks, note that pair 5 and 13 have similar sizes). The no template control (NTC) showed a different pattern of non-specific amplicons. M: molecular size marker. **d**, IGV plots showing Nanopore sequencing read coverage of the SARS-CoV-2 genome. All samples showed reads covering all of the targeted regions. **e**, Pipeline of RTNano real-time analysis. RTNano monitors the Nanopore MinION sequencing output folder. Once newly generated fastq files are detected, it moves the files to the analyzing folder and makes a new folder for each sample. If the Nanopore demultiplexing tool guppy is provided, RTNano will do additional demultiplexing to make sure reads are correctly classified. The analysis will align reads to the SARS-CoV-2 reference genome, filter, and count alignment records, and assign result mark (POS, NEG or UNK) for each sample. As sequencing proceeds, RTNano will merge the newly analyzed results with existing ones to update the current sequencing statistics.

## Results

Singleplex RPA was first performed to test its ability to amplify the SARS-CoV-2 genome from a nasopharyngeal swab sample tested positive for the virus using the US CDC assays^5^ (CT value=21). Sixteen primers were tested in 12 combinations to amplify 5 regions harboring either reported signature mutations^19,20^ useful for strain classification or mutation hotspots (GISAID, as of March 15, 2020). Five pairs of primers showed robust amplification of DNA of predicted size (range: 194-466 bp) in a 20-min isothermal reaction at 39 °C (Fig. 1b, Supplementary Fig. 1a). The specificity of all 5 RPA products was verified by restriction enzyme digestion (Supplementary Fig. 1b). The limit of detection of RPA reaches below 10 copies (Supplementary Fig. 1c). Furthermore, we showed that these RPA reactions could be multiplexed to amplify the five regions of the SARS-CoV-2 genome in a single reaction (Fig. 1c), thus significantly simplifying the workflow.

We next performed multiplex RPA using ten SARS-CoV-2 positive samples (SARS-CoV-2^+^, determined by US CDC assays^5^, CT value range: 15 to 27.9). Multiplex RPA products of the 10 samples were individually barcoded, pooled and prepared into a Nanopore sequencing library using an optimized protocol to save time. The whole workflow from RNA to sequencing took approximately 4 hours (Fig. 1a). The barcoded library was sequenced on a Nanopore MinION using a R9.4.1 flow cell. After 12-h sequencing, we acquired a total of 1.7 million reads from this barcoded library (Supplementary Fig. 2a). The demultiplexed reads were distributed relatively evenly among the barcodes (samples). We further aligned the reads to the SARS-CoV-2 reference genome and found that all RPA amplicons were covered by thousands of reads in all samples, suggesting that barcoded multiplexed RPA sequencing worked effectively (Fig. 1d, Supplementary Fig. 2b).

The identification of a SARS-CoV-2 positive sample can be achieved by surveying the existence of targeted amplicons via sequencing. However, the determination of negative samples needs to rule out potential sample collection failure, thus requiring a sample quality validation control. We used the existence of transcripts of the human housekeeping gene *ACTB* as a quality check of sample collection. The sequencing results showed that the *ACTB* gene could be effectively amplified without significantly affecting the amplification of SARS-CoV-2 (Supplementary Fig. 2c).

Co-infection with other respiratory pathogens in COVID-19 patients has been evaluated, however, current detection of co-infections requires additional rRT-PCR assays^9,21,22^ or NGS^8^. Such ad hoc tests are not amenable to large-scale screening of co-infections. On the other hand, multiplexed RPA of different viruses could be a promising solution to detect co-infections in a timely and efficient manner.

As a proof-of-principle of multiplex RPA of multiple human viral pathogens, we screened and validated RPA primers that robustly amplify influenza A (FluA), human adenovirus (HAdV), and non-SARS-CoV-2 human coronavirus (HCoV), respectively (data not shown). We added the three pairs of respiratory virus primers to the multiplex SARS-CoV-2 RPA panel to achieve simultaneous isothermal amplification of four viral pathogens (Supplementary table 1). To ensure effective amplification of all targets, we adjusted the concentration of each primer pair based on the read depth of the cognate genomic region when sequencing a contrived co-infection sample made from a SARS-CoV-2^+^ sample spiked with a control panel of 21 respiratory viruses including FluA, HAdV, and HCoV (Respiratory21^+^). The final primer mix achieved amplification of all targeted amplicons (Fig. 2a).

**Figure 2.**
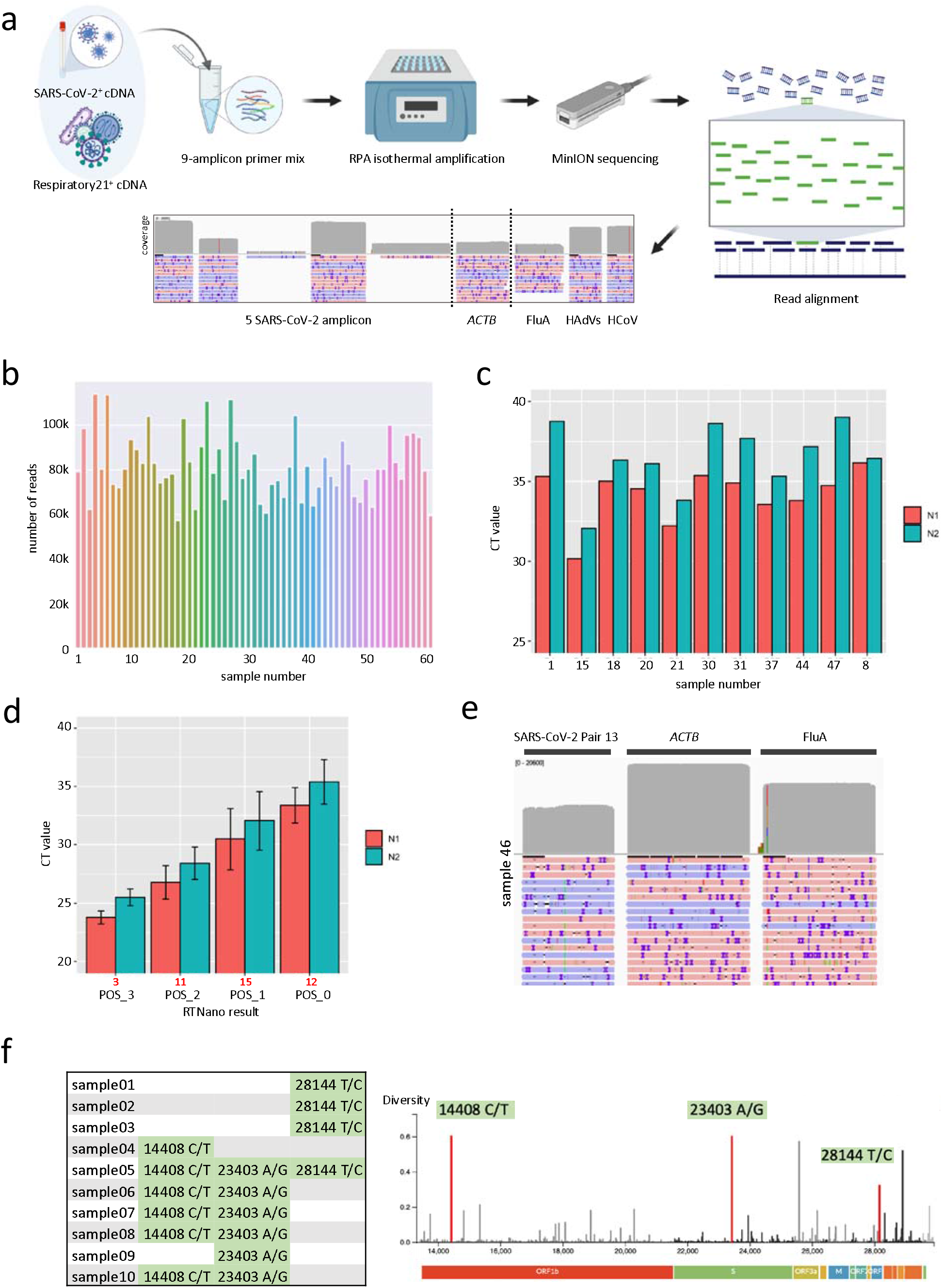
Real-time detection of multiple viral pathogens and mutational analysis of SARS-CoV-2. **a**, Experimental design of multiple virus detection by one-pot NIRVANA. A mixture of SARS-CoV-2^+^ and Respiratory21^+^ samples was used as positive control to adjust the primer concentration. The final primer mix could amplify all targeted viral regions. **b**, The sequencing throughput of 60 clinical samples (1-60) and NTC (61). A total of 6.3 million reads were acquired in a 24-hour sequencing run. **c**, CT values of potentially false-negative samples by RTNano analysis. The average CT value of the N1 assay was indicated by the blue line. **d**, The average rRT-PCR CT values of SARS-CoV-2 RTNano^+^ samples (PCR^+^ of both N1 and N2 assays) of different confidence level using 9-amplicon NIRVANA. The sample number is shown in red under the graph. RTNano confidence level inversely correlates with CT value. **e**, IGV plots showing the read alignment to the SARS-CoV-2, *ACTB*, and FluA amplicon in sample 46 using 9-amplicon NIRVANA. **f**, The SNVs detected in multiplex RPA sequencing and their position as shown in the Nextstrain data portal (Nextstrain.org). A total of 16 SNVs were detected from 10 SARS-CoV-2 positive samples.

The presence of SARS-CoV-2 has been reported in municipal sewage, of which the viral load was found to be correlated to the reported COVID-19 prevalence^23^. This suggests that wastewater surveillance could be a sensitive indicator of total COVID-19 case load (including asymptomatic cases) in the population. To evaluate the possibility of using multiplex RPA to detect SARS-CoV-2 in wastewater, we determined the primer mix to simultaneously amplify five regions of SARS-CoV-2 and one region of pepper mild mottle virus (PMMoV, an omnipresent indicator of water quality^24^). We spiked the wastewater concentrate (see methods) with RNA of SARS-CoV-2 positive samples, and used this as a positive control to test the multiplex RPA. Two primer mixes with different concentration of PMMoV primers were tested. Both primer mixtures can detect the SARS-CoV-2 and PMMoV within the positive control (Supplementary Fig. 2d). These results suggest that multiplex RPA can be used to monitor the presence of SARS-CoV-2 and other viruses in municipal wastewater. In addition, because the simultaneous acquisition of viral sequences, this method can also survey the strains circulating in the population (see below).

We next performed multi-virus multiplex RPA assay followed by Nanopore sequencing in 60 clinical samples suspected of SARS-CoV-2 infection. Following RT and RPA (Fig. 1a), amplicons of each sample were barcoded and sequenced in one Nanopore MinION flow cell. To take advantage of the unique feature of real-time base-calling offered by Nanopore sequencing, we developed a bioinformatics algorithm termed Real-Time Nanopore sequencing monitor (RTNano) (Fig. 1e). RTNano continuously monitors the output folder of basecalled data during the sequencing run and generates analysis reports in a matter of seconds. After detecting new fastq files (basecalled sequence output format of Nanopore sequencing), RTNano quickly aligns the reads to the targeted amplicons of the viruses. To confidently determined the existence of a virus, RTNano filters the alignment records by percentage identity and amplicon coverage and provides the number of positive records for each targeted viral amplicon, which is then used to evaluate the infection status of the sample.

When demultiplexing a large number of samples, barcode misclassification could potentially happen due to base-calling errors in the barcodes. RTNano includes an additional round of demultiplexing using stringent parameters to reduce the chance of barcode misclassification (see methods). Furthermore, a no template control (NTC) is included to monitor barcode misclassification. The reads of targeted amplicons assigned to the NTC barcode likely represented background errors in demultiplexing. The positive record number of the NTC will be subtracted from individual sample analysis to further reduce false positive identification. After the real-time analysis, RTNano generates a report for each of the 60 sample, including current read number, base number, and details of alignment positive records (Fig. 1e). To simplify the interpretation of the result, RTNano provides a summary score of SARS-CoV-2 based on predefined rules (Supplementary table 2). SARS-CoV-2 positive samples (POS) are assigned with different confidence levels based on the number of covered amplicons and corresponding positive records, range from 0 to 3 (lowest to highest). When there is no record of SARS-CoV-2, the sample could be categorized as either negative (NEG) if there are enough records of *ACTB*; or unknown (UKN) if there are insufficient records of *ACTB*. The introduction of confidence level can improve the accuracy of diagnosis, which is in principle similar to the CT value of rRT-PCR but based on quantitative information of multiple amplicons. Since base calling and demultiplexing may lead to a brief delay, we refer to the workflow as <u>N</u>anopore sequencing of <u>I</u>sothermal <u>R</u>apid <u>V</u>iral <u>A</u>mplification for <u>N</u>ear real-time <u>A</u>nalysis (NIRVANA) (Fig. 1a).

RTNano identified 35 SARS-CoV-2 positive cases in the 60 samples after 2 hours of Nanopore sequencing. Ten more positives were identified by RTNano (RTNano^+^) as data output increased in the next 22 hours of sequencing (Fig. 2b, Supplementary File 1). An identical aliquot of each sample was analyzed in parallel using the US CDC rRT-PCR SARS-CoV-2 assays, 57 samples were confirmed as SARS-CoV-2 positive or negative, while 3 samples were inconclusive after two rounds of tests (Supplementary File 2). We excluded the 3 inconclusive samples in the downstream analysis. Among the analyzed 43 RTNano^+^ samples, 41 (95.35%) were also considered positive by the rRT-PCR assay (PCR^+^). The two remaining weakly RTNano^+^ sample (POS_0) had high CT values of RNase P and failed in PCR amplification of N1 and N2. Three of the RTNano^-^ samples were also PCR^-^ due to failed amplification of N1 and/or N2, while the rest of 11 RTNano^-^ were PCR^+^. These 11 false-negative samples had high CT values (N1: 34.16 ± 0.36, N2: 36.27 ± 0.47, Fig. 2c). The incorrect identification of these samples could be due to amplification failure of multiplex RPA or insufficient sequencing throughput. In addition, high CT values in SARS-CoV-2 rRT-PCR test have a higher chance of being false-positive^25,26^.

We calculated the average CT value of RTNano^+^ samples under different confidence levels. RTNano^+^ samples with high confidence level correlated with lower CT values, which proved the reliability of RTNano results (Fig. 2d). Based on the N1 standard curve, we estimated the limit of detection (LoD) of NIRVANA was ∼29 viral RNA copies/µl of extracted nucleic acid (86 copies per reaction, based on average N1 CT value of 33.37 of POS_0, and see methods), which was comparable to the current rRT-PCR assays^27^.

We next surveyed co-infection of three common respiratory viruses in the 60 samples. The ground truth of the FluA co-infection was determined by both in vitro diagnostic kit (CE-IVD) Resp’Easy^™^ and IDT Influenza/RSV primers. Four samples were identified as FluA^+^ in both assays with CT values from 33.97 to 39.33 (by Resp’Easy^™^) (Supplementary Fig. 2e). RTNano detected the FluA^+^ sample with the lowest CT value (sample 46, CT 33.97) with high confidence (Fig. 2e). The HAdVs and HCoV co-infection was examined using the Resp’Easy^™^ kit, and only one HCoV^+^ sample was identified (Resp’Easy^™^ CT=35.18). RTNano reported no HAdVs co-infection, while failed to detect the HCoV^+^ sample.

A high level of multiplexing reactions could negatively affect RPA efficiency of each amplicon. To test if a less complex primer combination could improve the performance of NIRVANA, we removed less robust SARS-CoV-2 primers (pair 9 & 10) and performed a new NIRVANA sequencing. Twenty-nine of the 60 samples were sequenced, including 11 RTNano^-^/PCR^+^, 4 FluA^+^, and 1 HCoV^+^. Nine of the previous 11 RTNano^-^/PCR^+^ samples were identified as SARS-CoV-2^+^, suggesting an improved sensitivity (Supplementary File 3). The average N1 CT value of POS_1 was improved from 30.48 to 32.06 (Supplementary Fig. 2f). Using the new data, we calculated the LoD of SARS-CoV-2 of 7-amplicon NIRVANA was ∼20 viral RNA copies/µl of extracted nucleic acid (61 copies per reaction, based on average N1 CT value of 33.37 of POS_0).

The sensitivity of FluA detection was also improved as two FluA^+^ samples (Resp’Easy^™^ CT value of 33.97 and 36.03) were correctly identified (sample 46 & 58 in Supplementary Fig. 2e, Supplementary Fig. 2g). The HCoV^+^ sample was still not identified. Since NIRVANA showed a robust ability in detecting HCoV in the positive control sample (Fig. 2a), we suspected that the HCoV in the clinical sample may belong to a different strain that could not be amplified by our HCoV primers. Taken all together, these results showed that NIRVANA provided high-confident SARS-CoV-2 detection for virus loads above 20 copies/µl of extracted nucleic acid and reliable capability in detecting potential co-infections.

RTNano has an integrated function to quickly analyze variants in each sample during sequencing. We used it to analyze sequence variants in ten SARS-CoV-2 positive samples (Fig. 1d). It detected 16 single nucleotide variants (SNVs) in the ten samples and all of them had been reported in GISAID (Fig. 2f, as of June 7, 2020). The reported SNVs suggested that the strains in samples 01-03 are close to clade 19B (nt28144 T/C), first identified in Wuhan, China, while the strains in samples 04-10 are close to clade 20 (nt14408 C/T, 23403 A/G), first becoming endemic in Europe. Given the fact that samples 01-03 were collected early in the pandemic, while the others were later (around May), the SNV signature of the strains revealed by NIRVANA is consistent with the pattern of COVID-19 case importation, which initially came from China and shifted to Europe after a ban of flight from China. Prospectively collecting such data regularly could guide public health policy making to better control the pandemic.

To validate the SNVs and compare NIRVANA with conventional RT-PCR amplicon sequencing^28^, we chose three samples (01-03) to perform multiplex RT-PCR amplicon sequencing on the Nanopore MinION. Variant calling was done by RTNano using the same parameters and the results showed that RT-PCR sequencing confirmed all 3 SNVs detected by NIRVANA (Supplementary Fig. 3a). We further compared the SNVs of samples 01-03 with their corresponding assembled genome from Illumina sequencing published in GISAID (EPI_ISL_437459 for sample03, EPI_ISL_437460 for sample04, and EPI_ISL_437461 for sample05). All of the three SNVs existed in the assembled genome.

Taken together, NIRVANA provides high-confidence detection of both SARS-CoV-2 and other respiratory viruses, and mutation surveillance of SARS-CoV-2 on the fly. Compared to the Oxford Nanopore’s official method termed LAMPore^29^, which is based on RT-LAMP, NIRVANA offers several advantages including longer amplicons and higher level of multiplexing. LAMPore generated short amplicons (∼80 bp), of which half was composed of primer sequences. Thus, the read alignment may be more prone to amplification artifacts that lead to false-positive calling. In NIRVANA, significantly longer amplicons (up to 466 bp in this study) combined with alignment record filter can significantly improve the accuracy of positive result by eliminating the influence of amplification artifacts. In addition, LAMP-based LAMPore requires multiple primers (at least 4) to amplify one amplicon, which was challenging for multiplexing. In contrast, we showed that RPA-based NIRVANA is able to amplify and detect nine amplicons in one pot (Fig. 2a). It holds great promise in detecting multiple viruses simultaneously to help monitor and control the pandemic and the treatment of patients.

The whole workflow of NIRVANA can be further shortened by performing one-pot RT-RPA (Supplementary Fig. 1d) so that the time from RNA to answer can be as short as 3.5 hours. All molecular biology reactions in the workflow can be done in a simple heating block, and all necessary supplies fit into a briefcase (Supplementary Fig. 3b). The framework of NIRVANA was built with flexibility of detecting other viruses by changing primers. We expect it to provide a rapid field-deployable solution of pathogen detection and surveillance of pandemic strains.

## METHODS

### RNA samples and primers

Anonymized RNA samples were obtained from Ministry of Health (MOH) hospitals in the western region in Saudi Arabia. The use of clinical samples in this study is approved by the institutional review board (IRB# H-02-K-076-0320-279) of MOH and KAUST Institutional Biosafety and Bioethics Committee (IBEC). Oropharyngeal and nasopharyngeal swabs were carried out by physicians and samples were steeped in 1 mL of TRIzol (Invitrogen Cat. No 15596018) to inactivate virus during transportation. Total RNA extraction of the samples was performed following instructions as described in the CDC EUA-approved protocol using the Direct-Zol RNA Miniprep kit (Zymo Research Cat. No R2070) or TRIzol reagent (Invitrogen Cat. No 15596026) following the manufacturers’ instructions. The Respiratory (21 targets) control panel (Microbiologics Cat. No 8217) was used as positive control in the amplification of FluA, HAdVs and HCoV. A list of primers used in this study can be found in Supplementary table 1.

### Wastewater samples

Raw sewage was collected at 9 AM and 4 PM on 7 June 2020 from the wastewater equalization tank in KAUST, and then mixed together to constitute a composite sample. A 300-500 ml of sewage mixture was concentrated by electronegative membrane in the present of cation as previously described^30^. The eluate of viruses was recovered in a tube with 50 μL of 100 mM H_2_SO_4_ (pH 1.0) and 100 μL of 100× Tris– EDTA buffer (pH 8.0) for neutralization. Centripep YM-50 (Merck Millipore) was used to further concentrate the samples to a volume of 600-700 µl.

### Reverse transcription

Reverse transcription of RNA samples was done using either NEB ProtoScript II reverse transcriptase (NEB Cat. No M0368) or Invitrogen SuperScript IV reverse transcriptase (Thermo Fisher Scientific Cat. No 18090010), following protocols provided by the manufacturers. After reverse transcription, 5 units of thermostable RNase H (New England Biolabs Cat. No M0523S) was added to the reaction, which was incubated at 37 °C for 20 min to remove RNA. The final reaction was diluted to be used as templates in RPA. All of the web-lab experiments in this study were conducted in a horizontal flow clean bench to prevent contaminations. The bench was decontaminated with 70% ethanol, DNA*Zap* (Invitrogen, Cat no. AM9890) and RNase *AWAY* (Invitrogen, Cat no. 10328011) before and after use. The filtered pipette tips (Eppendorf epT.I.P.S.® LoRetention series) and centrifuge tubes (Eppendorf DNA LoBind Tubes, Cat. No 0030108051) used in this study were PCR-clean grade. All of the operations were performed carefully following standard laboratory operating procedures.

### rRT-PCR

The rRT-PCR assay of SARS-CoV-2 was purchased from IDT (Cat. No 10006770). The rRT-PCR assay for FluA was purchase from IDT (Cat. No 1079729). Respiratory Virus PCR Panel kit (Diagenode diagnostics, DDGR-90-L048) was used for the determination of FluA, HAdVs and HCoV. All reactions were run on a CFX384 Touch Real-Time PCR Detection System (Bio-rad) following the instruction of manufacturers. The copy number of SARS-CoV-2 is determined by the IDT SARS-CoV-2 rRT-PCR assay based on the standard curve of CT values of known copies of synthetic SARS-CoV-2 N gene RNA (nt28,287-29,230 in NC_045512.2, DNA template purchased from IDT). A serial dilution of the synthetic RNA is performed to obtain final concentrations of 10, 10^2^, 10^3^, 10^4^, 10^5^, 10^6^, 10^7^ copies/µl. The reverse transcription is done using 1 µl of diluted RNA, followed by rRT-PCR using 2.5 µl of 5-fold diluted cDNA.

### Singleplex RPA

Singleplex RPA was performed using TwistAmp® Basic kit following the standard protocol. Thirteen pairs of SARS-CoV-2 primers covering N gene, S gene, ORF1ab and ORF8 were tested and the corresponding amplicons were purified by 0.8X Beckman Coulter AMPure XP beads (Cat. No A63882) and eluted in 40 µl H_2_O. The purified amplicons were first analyzed by running DNA agarose gel to check the specificity and efficiency. The most robust five pairs of primers with correct size were further analyzed by NlaIII (NEB Cat. No R0125L) and SpeI (NEB Cat. No R0133L) digestion following standard protocols. For one-pot RT-RPA, 10 U of AMV Reverse Transcriptase (NEB Cat no. M0277S) and 20 U of SUPERase•In™ RNase Inhibitor (Invitrogen Cat no. AM2694) were added to a regular RPA reaction mix. The RT-RPA was carried out per the manufacturer protocol. For the RPA of respiratory viruses, reverse-transcribed cDNA from the Respiratory (21 targets) control panel (Microbiologics Cat. No 8217) was used as the template.

### Multiplex RPA

Multiplex RPA was done by add all of the primers in the same reaction. The total final primer concentration is set to 2 μM. To achieve an even and robust amplification, we empirically determined the final concentration of the five-amplicon primers (for the initial test of 10 SARS-CoV-2 positive samples) as follows: 0.166 µM for each pair-4 primer, 0.166 µM for each pair-5 primer, 0.242 µM for each pair-9 primer, 0.26 µM for each pair-10 primer, 0.166 µM for each pair-13 primer, 29.5 µl of primer free rehydration buffer, 1 µl of 10-fold diluted cDNA, 7µl H_2_O. In the multiplex RPA of 9-amplicon and 7-amplicon (remove pair 9 & 10) primers, the primer mixtures were obtained by combining different amount of 10 µM primers according to the ratios in Supplementary table 1, and 2.5 µl of 5-fold diluted cDNA were used as template. The reaction was incubated at 39 °C for 4 min, then vortexed and spin down briefly, followed by a 16-min incubation at 39 °C.

### Library preparation and sequencing

The RPA library preparation was done using Native barcoding expansion kit (Oxford Nanopore Technologies EXP-NBD114 and EXP-NBD196) following Nanopore PCR tiling of COVID-19 Virus protocol (Ver: PTC_9096_v109_revE_06Feb2020) with a few changes to save time. The RPA reaction was purified using Qiagen QIAquick PCR purification kit (Qiagen Cat No. 28106) and elute in 30 µl H_2_O. The end-prep reaction was done separately in 15 µl volume using 5 µl of each multiplex RPA samples. After that, we followed the same procedures as described in the official protocol. The RT-PCR library preparation was done using Native barcoding expansion kit (Oxford Nanopore Technologies EXP-NBD104) according to the standard native barcoding amplicons protocol. The sequencing runs were performed on an Oxford Nanopore MinION sequencer using R9.4.1 flow cells.

### Bioinformatics

RTNano scanned the sequencing folder repeatedly based on user defined interval time. Once newly generated fastq files were detected, it moved the files to the analysis folder and made a new folder for each sample. If the Nanopore demultiplexing tool guppy is provided, RTNano will do additional demultiplexing to make sure reads are correctly classified. The analysis part utilized minimap2^31^ to quickly align the reads to the SARS-CoV-2 reference genome (GenBank: NC_045512). After alignment, RTNano will filter the alignment records based on defined thresholds of parentage identity and amplicon coverage, followed by counting the alignment records of each amplicon. A read with >= 89% alignment identity and >= 96% amplicon coverage will be counted as one positive record. If an NTC barcode number was provided, RTNano will subtract this number in individual sample analysis to further ensure confident demultiplexing. In the end, RTNano will assign samples with different result marks (POS, NEG and UNK) based on the number of alignment records of each amplicon. With the sequencing continuing, RTNano will merge the newly analyzed result with completed ones to update the current sequencing statistics. RTNano is ultra-fast, a typical analysis with additional guppy demultiplexing of 5 fastq files (containing 4000 reads each and sequenced with 12-barcode kit) will take ∼10s using one thread in a MacBook Pro 2016 15-inch laptop. Variant calling was performed using samtools (v1.9) and bcftools (v1.9)^32^. The detected variants were filtered by position (within the targeted regions) and compared with the data in Nextstrain.org as of Jun 2, 2020.

### Data and materials availability

RTNano and sample data in this study are accessible at GitHub (https://github.com/milesjor/RTNano).

## Data Availability

RTNano and sequencing data in this study are available upon request.  

## Acknowledgements

We thank members of the Li laboratory, Baolei Yuan, Xuan Zhou, Yingzi Zhang, and Samhan Alsolami for helpful discussions; Marie Krenz Y. Sicat for administrative support. We thank members of the Pain lab, Rahul Salunke and Amit Subudhi for technical assistance. We thank members of the Izpisua Belmonte lab, Yanjiao Shao, Yasuo Ouchi, and Pradeep Reddy for generously sharing ideas of simultaneous pathogen detection. We thank Professor Peiying Hong and her lab members, Dr. Andri Rachmadi and Dr. David Mantilla-Calderon for kindly sharing wastewater samples and technical assistance.

## Author contributions

ML and CB performed majority of the experiments related to Nanopore sequencing. GM and CB performed diagnosis assays for various viral pathogens. CB wrote the code and performed the bioinformatics analysis. ML, CB, JX and CRE designed and performed molecular biology experiments. FSA, AK, AMH, and NAMA collected clinical samples. SM extracted RNA and performed molecular assays. SH and AP coordinated the clinical samples and molecular testing. CB, GM, YT, END, JCBI and ML analyzed the data and wrote the manuscript. ML, CB and JCBI conceived the study. JCBI and ML supervised the study.

## Supplementary Figures

**Supplementary Figure 1.**
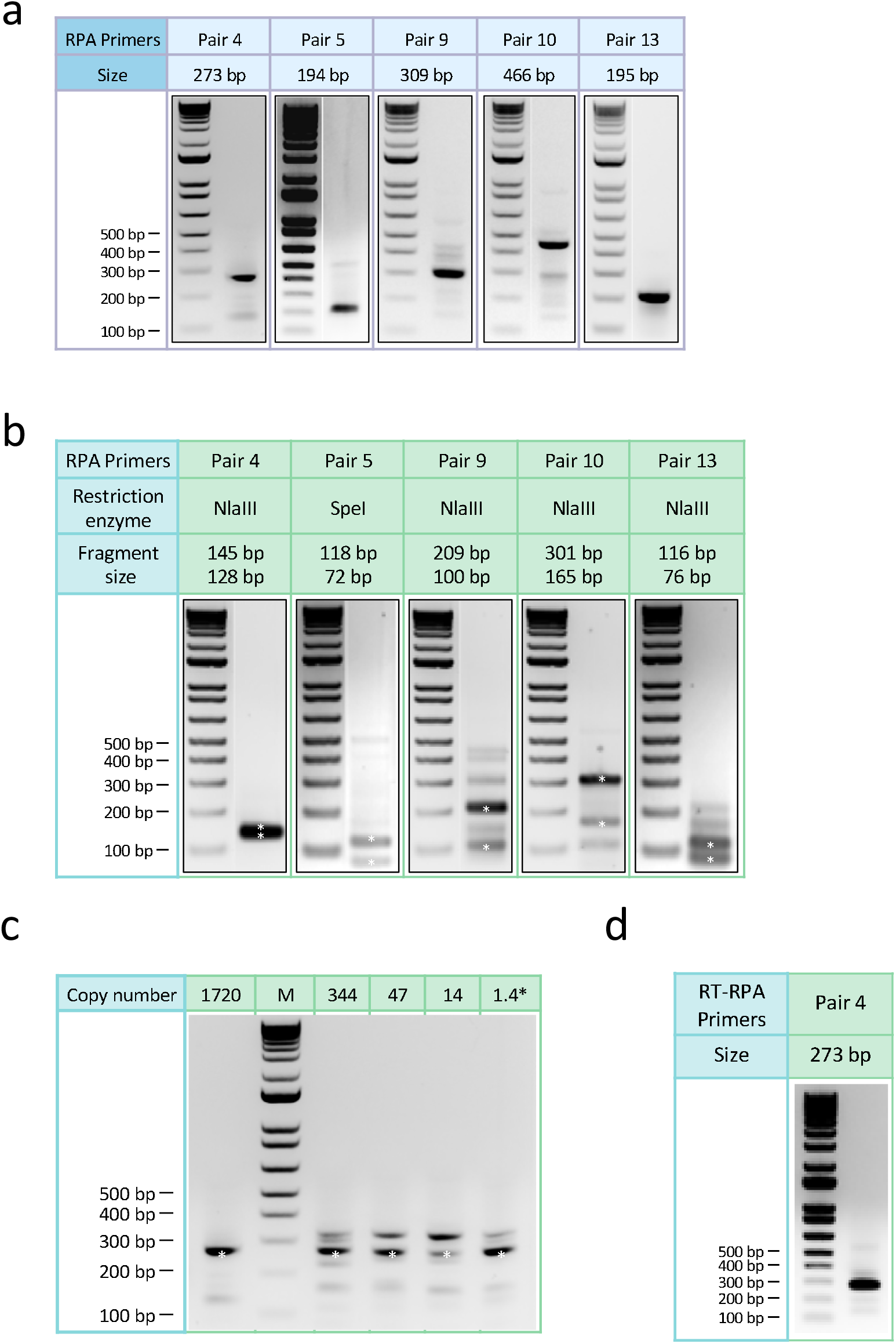
Agarose gel electrophoresis results of singleplex RPA. **a**, Agarose gel electrophoresis results of singleplex RPA with selected primers shown next a molecular size marker. The amplicons range from 194 bp to 466 bp. **b**, Agarose gel electrophoresis results of restriction enzyme digestion. The amplicon of pair 5 was digested by SpeI while the others were digested by NlaIII. The digested DNA bands (asterisks) were of expected sizes. **c**, Agarose gel electrophoresis results showing the sensitivity of RPA in amplifying the SARS-CoV-2 genome. Primer pair 4 was used in the experiment. Reliable amplification can be achieved with 1.4 copies (calculated from dilution) of the SARS-CoV-2 genome. **d**, Agarose gel electrophoresis result of one-pot reverse transcription and RPA reaction using primer pair 4.

**Supplementary Figure 2.**
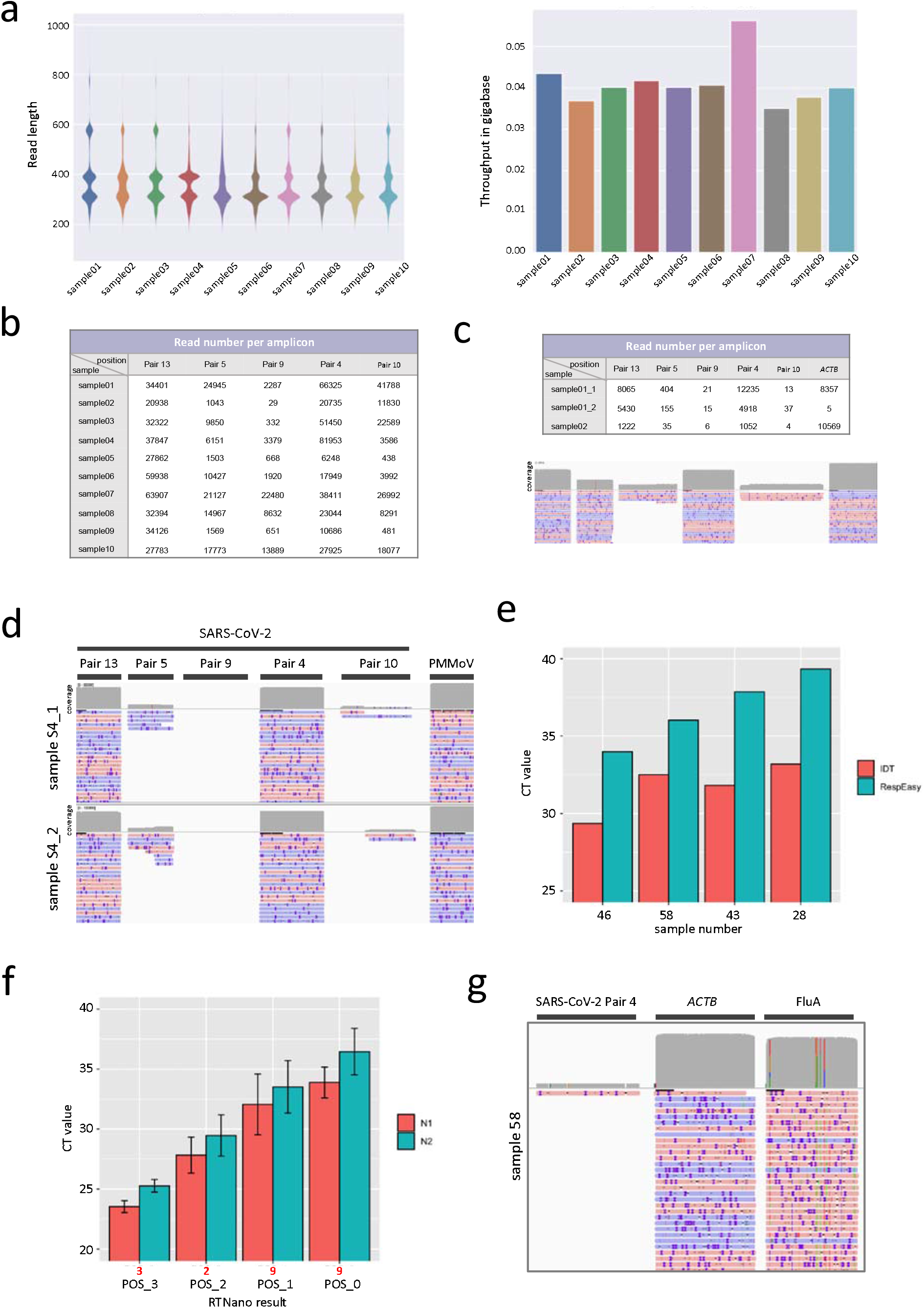
Sequencing analysis of multiplex RPA. **a**, The read length distribution and throughput of ten SAR-CoV-2^+^ sample sequencing. **b**, The read number for each amplicon in the sequencing of ten SAR-CoV-2^+^ samples. All amplicons were covered by reads. **c**, The read number for each amplicon in the trial sequencing of multiplex RPA of SARS-CoV-2 and *ACTB*. Sample 01 is used as RPA template to determine the primer concentration in two trials (sample01_1 and sample01_2). Sample 02 is used in a repeat trial using the same primer mix as sample01_2. **d**, The IGV alignment plot showing robust amplification of PMMoV with SARS-CoV-2. A SARS-CoV-2^+^ sample (S4) was used as input sample in two trials with different primer concentration. **e**, The CT values of FluA^+^ samples in Resp’Easy^™^ and IDT FluA assays. **f**, The average rRT-PCR CT values of SARS-CoV-2 RTNano^+^ samples (PCR^+^ of both N1 and N2 primers) of different confidence level using 7-amplicon NIRVANA. **g**, IGV plots showing the read alignment to SARS-CoV-2, *ACTB* and FluA amplicon in sample 58 using 7-amplicon NIRVANA.

**Supplementary Figure 3.**
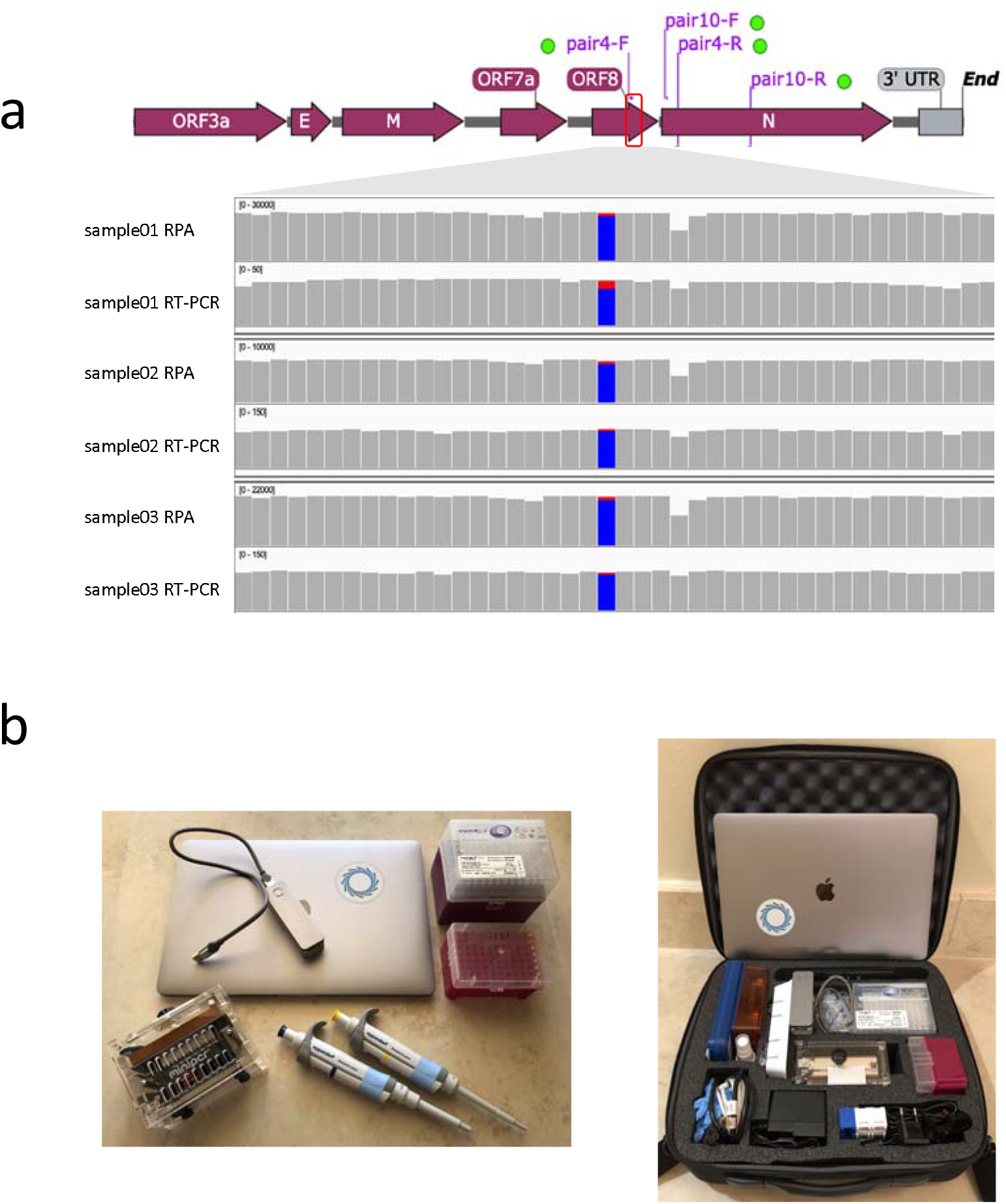
Validation of SNVs detected by NIRVANA. **a**, IGV plots showing the nt28144 T/C SNV in samples 01-03 from RPA and RT-PCR Nanopore sequencing. The blue bar represents the C base while the red bar represents the T base. All of the 3 SNVs detected in RPA sequencing were confirmed by RT-PCR amplicon sequencing. **b**, Equipment used in NIRVANA. The whole workflow can be done with one laptop, one Nanopore MinION sequencer, two pipettes, two boxes of pipette tips, and a heating block (using a miniPCR™ mini16 here). All equipment can be packed into a suitcase.

